# Different patterns of structural network impairments in two amyotrophic lateral sclerosis subtypes driven by ^18^F-FDG PET/MR hybrid imaging

**DOI:** 10.1101/2023.11.02.23297955

**Authors:** Feng Feng, Guozheng Feng, Jiajin Liu, Weijun Hao, Weijie Huang, Xiao Bi, Mao Li, Hongfen Wang, Fei Yang, Zhengqing He, Jiongming Bai, Haoran Wang, Guolin Ma, Baixuan Xu, Ni Shu, Xusheng Huang

**Author notes:** **Corresponding Authors** Baixuan Xu, Department of Nuclear Medicine, First Medical Center, Chinese PLA General Hospital, No.28, Fuxing Road, Haidian District, Beijing, 100853, China, Ni Shu, Beijing Key Laboratory of Brain Imaging and Connectomics, Beijing Normal University, No.19, Xinjiekouwai Street, Haidian District, Beijing, 100875, China, Xusheng Huang, Department of Neurology, First Medical Center, Chinese PLA General Hospital, No.28, Fuxing Road, Haidian District, Beijing, 100853, China. These authors have contributed equally to this work (Feng Feng, Guozheng Feng, Jiajin Liu).

## Abstract

The structural network damages in amyotrophic lateral sclerosis (ALS) patients are evident but contradictory due to the high heterogeneity of disease. We hypothesized that patterns of structural network impairments would be different in ALS subtypes by a data-driven method using ^18^F-FDG PET/MR hybrid imaging. 50 patients with ALS and 23 healthy controls (HCs) were collected PET, structural MRI and diffusion tensor imaging data by a ^18^F-FDG PET/MR hybrid. Two ALS subtypes were identified as the optimal cluster based on gray matter volume and standardized uptake value ratio. Network metrics at the global, local and connection levels were compared to explore the impaired patterns of structural network in the identified subtypes. Compared with HCs, the two ALS subtypes displayed a pattern of a locally impaired structural network centralized in the sensorimotor network and a pattern of an extensively impaired structural network in the whole brain. When comparing the two ALS subgroups by a support vector machine classifier based on the decreases in nodal efficiency of structural network, the individualized network scores were obtained in every ALS patient and demonstrated a positive correlation with disease severity. We clustered two ALS subtypes by a data-driven method, which encompassed different patterns of structural network impairments. Our results imply that ALS may possess the intrinsic damaged pattern of white matter network and thus provide a latent direction for stratification in clinical research.

## 1. Introduction

Amyotrophic lateral sclerosis (ALS) is a progressive neurodegenerative disease that mainly involves upper and lower motor neurons. Although muscle weakness, atrophy and fasciculation are the most predominant symptoms of ALS, the high heterogeneity in clinical manifestations is also remarkable. ALS can be categorized into familial and sporadic subtypes, bulbar-onset and limb-onset subtypes, as well as fast-progression, intermediate-progression and slow-progression subtypes (van Es et al., 2017). On the basis of cognition level, ALS can be divided into ALS with normal cognition, ALS with cognitive impairment (ALS-ci), ALS with behavioral impairment (ALS-bi), ALS with cognitive and behavioral impairment (ALS-cbi), and ALS with frontotemporal dementia (ALS-FTD) (Strong et al., 2017). Taken together, these classifications are all based on clinical characteristics.

Recently, cluster analysis, as a data-driven method, has been used to identify subtypes of Alzheimer’s disease (Noh et al., 2014; Ten Kate et al., 2018) and behavioral variants of FTD (Whitwell et al., 2009) with neuroimaging data. However, cluster analysis with MRI and PET data has not been used in ALS patients to date. In ALS patients compared to healthy controls (HCs), reductions in gray matter volume (GMV) in the bilateral precentral gyri were observed using FreeSurfer software with structural MRI (sMRI) data (Kwan et al., 2012), and decreases in the standardized uptake value ratio (SUVR) in the frontal, motor, and parietal cortices were demonstrated by PET with ^18^F-fluorodeoxyglucose (FDG) (Matías-Guiu et al., 2016; Pagani et al., 2014; Sala et al., 2019). Considering that a single-modal metric provides limited information, a combination of GMV and SUVR was hypothesized to be a complex data-driven marker that could reflect the characteristics of disease in more detail. Although sMRI and ^18^F-FDG PET have been combined to collect neuroimaging data in ALS patients (Buhour et al., 2017), these data were acquired at different times and thus inevitably with errors of image registration.

Fortunately, with the advantage of simultaneous data collection, PET/MR hybrid scans are not subject to registration errors and can achieve the integration of multimodal metrics. In previous studies of ALS patients using ^11^C-PBR28, the imaging data were collected by an integrated PET and MRI system, which was not a PET/MR hybrid in the true sense (Alshikho et al., 2016; Ratai et al., 2018). In ALS patients, PET/MR hybrid scans using ^18^F-DPA714 and ^11^C-JNJ717 have been reported (Van Weehaeghe et al., 2020). In patients with ALS or behavioral variants of FTD plus motoneuron disease, a significant increment in glucose metabolism in the midbrain/pons and medulla oblongata was found in comparison to controls by ^18^F-FDG PET/MR (Zanovello et al., 2022).

Due to the remarkable involvement of motor neurons, previous studies on brain connectivity in ALS patients mainly focused on structural connectivity with diffusion tensor imaging (DTI) data. By graph theory, previous observations have exhibited decreases in the global efficiency of structural network (Fortanier et al., 2019; Zhang et al., 2019) and in nodal topological centralities in the frontal, parietal and temporal lobes when comparing ALS patients to controls (Li et al., 2021). By network-based statistics (NBS), an impaired structural subnetwork was revealed in ALS patients with a typical involvement of primary and secondary motor connections (Buchanan et al., 2015; Verstraete et al., 2011). Furthermore, ALS patients with bulbar onset and spinal onset both showed the most severely damaged connections mainly involving bilateral precentral gyri and paracentral lobules, but the spinal-onset group displayed a more widespread pattern of affected connections compared to controls (van der Burgh et al., 2020). However, ALS patients with different disease durations showed consistent involvement of the motor network and limited extramotor involvement (van der Burgh et al., 2020). Although the extension of structural connectivity damage alone is already known to be correlated with measures of disease severity in ALS, it is still necessary to explore the different patterns of structural network impairments in the data-driven subtypes for the high heterogeneity in clinical manifestations in ALS. Due to by the data-driven method not by the clinical features, exploring the impaired pattern of structural network in the data-driven subtypes could be a latent facilitation for stratified therapy in ALS.

Here, using ^18^F-FDG PET/MR hybrid data, we hypothesized that ALS subtypes can be identified by cluster analysis based on GMV and SUVR. Next, we assessed the individual patterns of impaired structural network in the identified ALS subtypes by graph theory at the global, local and connection levels. Furthermore, we observed changes in GMV and ^18^F-FDG metabolism in the clustered subtypes for a deep evaluation of the white matter (WM) connectome. Finally, we explored potential biomarkers for the phenotypes of ALS based on significantly different metrics of structural network.

## 2. Methods

### 2.1 Participants

Patients with clinically definite (36), probable (10) or possible (4) ALS according to the revised El Escorial (Brooks et al., 2000) (50; 28 men and 22 women; mean age at symptom onset 49.70 ± 8.68 years; mean age at PET/MR scan 51.10 ± 8.98 years) were recruited from Chinese PLA Hospital from July 1, 2020 to April 30, 2022. With the exception of 1 patient with a family history of ALS, all other patients had no family history of ALS or FTD. Among the 40 ALS patients who accepted genetic detection with consent, all had normal number of GGGGCC repeat expansions in the *C9orf72* gene and 33 displayed negative results of whole-exome sequencing. The 7 ALS patients with missense mutations were showed in Table S1. Furthermore, age-, sex- and education level-matched HCs (23; 10 men and 13 women; mean age at PET/MR scan 48.87 ± 10.81 years) were enrolled. All individuals or their legal guardians signed informed consent forms. All the participants were Han Chinese, right-handed, younger than 75 years old, without other neurological or psychiatric diseases, and without contraindications for PET and MRI examination.

### 2.2 Clinical assessments

All ALS patients and HCs were evaluated at length before the PET/MR scan. Dysfunction was measured with the ALS Functional Rating Scale-Revised (ALSFRS-R) (Cedarbaum et al., 1999). The progression rate from disease-onset to baseline (DeltaFS) was calculated using the following formula: 48 – (total ALSFRS-R at initial visit) /symptom duration (months) (Kimura et al., 2006). Fast, intermediate and slow ALS progressors were defined as DeltaFS ≥ 1.0, DeltaFS < 1.0 ∼ ≥ 0.5, and DeltaFS < 0.5, respectively (Lu et al., 2015; Zhang et al., 2022). The neuropsychological evaluations included the Mini-Mental State Examination (MMSE), Montreal Cognitive Assessment (MoCA), and Edinburgh Cognitive and Behavioral ALS Screen (ECAS) Chinese version (Ye et al., 2016). On the basis of the revised Strong criteria (Strong et al., 2017), ALS patients were diagnosed with normal cognition (28), ALS-ci (8), ALS-bi (5), ALS-cbi (4), and ALS-FTD (5). In the current study, ALS patients with normal cognition were called ALS-cn, and ALS-ci, ALS-bi, ALS-cbi and ALS-FTD were called ALS-plus. Two senior neurologists completed all the neuropsychological evaluations.

### 2.3 PET/MR scan

PET/MR scans with ^18^F-FDG were carried out by two senior nuclear medicine technicians in the Department of Nuclear Medicine, First Medical Center, Chinese PLA General Hospital, using a PET/MR hybrid scanner (Siemens, Biograph mMR). Before the PET/MR scan, T2-weighted images were collected to exclude the subjects with brain lesions. For the included participants, at least 6 hours of fasting and 30 minutes of rest in a quiet and dark environment were required before the intravenous injection of ^18^F-FDG (4.44–5.55 MBq/kg). Fifty minutes after the injection, a PET/MR scan was carried out. During the scan, a 16-channel head coil was used, and foam padding minimized head motion. The participants were asked to remain relaxed and keep their eyes open without falling asleep. ^18^F-FDG PET data were collected with the List model and further reconstructed by Poisson-ordered subset expectation-maximization algorithms with three iterations. Twenty-one subsets were obtained using a Gaussian filter of 2 mm full-width at half-maxima (FWHM) and 344 × 344 voxels. Next, DTI data were collected using a single-shot echo planar imaging (EPI) sequence in the axial plane. The EPI parameters were as follows: repetition time (TR) = 9,900 ms, echo time (TE) = 91 ms, acquisition matrix = 128 × 128, field of view (FOV) = 256 mm × 256 mm, and slice thickness = 2 mm with no gaps. A total of 70 contiguous slices were acquired for b values of 0 and 1,000 s/mm^2^ using gradients along 30 different diffusion directions. Finally, high-resolution sMRI data were obtained from sagittal T1-weighted images (T1WI; 192 continuous slices), which were acquired by a magnetization-prepared rapid gradient echo (MPRGE) sequence with the following scan parameters: TR = 1,900 ms, TE = 2.43 ms, inversion time (TI) = 1,100 ms, FOV = 256 mm × 256 mm, acquisition matrix = 512 × 512, flip angle = 9°, and slice thickness = 1 mm with no gaps.

### 2.4 Image processing

To obtain the GMV at cortical vertices in each hemisphere, T1WI was processed through the recon-all command in the FreeSurfer software package (https://surfer.nmr.mgh.harvard.edu/fswiki/FreeSurferWiki). Then, the Brainnetome Atlas (BNA) 246 (Fan et al., 2016) with 210 cortical regions and 36 subcortical regions was projected on native fsaverage to obtain the statistical GMV in each cerebral region according to the official scripts (http://www.brainnetome.org/resource/). Finally, the GMV in each cerebral region was divided by the mean GMV across all cerebral regions to obtain a normalized value.

To calculate the SUVR in the cerebral regions, T1WI for each subject was aligned to ^18^F-FDG PET images. Then, the aligned T1WI was transformed into the ICBM152 template in Montreal Neurological Institute (MNI) space using the FMRIB Linear Image Registration Tool (FLIRT) (https://fsl.fmrib.ox.ac.uk/fsl/fslwiki/FLIRT) (Jenkinson and Smith, 2001) and FMRIB Nonlinear Image Registration Tool (FNIRT) (https://fsl.fmrib.ox.ac.uk/fsl/fslwiki/FNIRT). These derived transformation matrices were applied to the BNA246 and automated anatomical labeling (AAL) 90 (Tzourio-Mazoyer et al., 2002) to obtain cerebral parcellations in native space. After smoothing using a 2 mm kernel on ^18^F-FDG PET images, the SUVR of each cerebral region was normalized to the mean SUVR across all cerebral regions.

Regarding DTI, preprocessing procedures comprised correction of eddy current and motion artifacts, diffusion tensor estimation, and fractional anisotropy (FA) calculation. Specifically, an affine alignment of each DTI to the b0 image was applied to correct eddy current distortions and motion artifacts using the eddy_correct command in the FDT toolbox of FSL (https://fsl.fmrib.ox.ac.uk/fsl/fslwiki/FDT). The diffusion tensor estimation and FA calculation were performed with the dtifit command in the FDT toolbox of FSL (Fan et al., 2016; Jenkinson and Smith, 2001).

### 2.5 Cluster analysis

As a data-driven clustering approach, nonnegative matrix factorization (NMF) can explore clusters of features in participants by an unsupervised strategy and was currently adopted to reveal ALS subtypes on NMF (v.0.23.0) in R (v.4.1.2) (Gaujoux and Seoighe, 2010). Features of each participant were characterized by the sum of SUVR and GMV values in the same cerebral region (Fig. 1a). Notably, the factors of age at PET/MR scan, sex ratio, and education years were removed by regression analysis before the calculation of GMV and SUVR in the cerebral regions. Next, the obtained GMV and SUVR values were further normalized by the min-max scaling method. ALS patients were clustered into different numbers of subtypes (cluster_n = 2, 3, …, 10) with various cophenetic correlation coefficients by similar inherent features. Based on the best fit (i.e., highest value of the cophenetic correlation coefficient), the optimal cluster (cluster_n = 2) was determined (Fig. 1b). With the optimal cluster, nonsignificant features by NMF were deleted from the feature selection (Fig. 1c). Using the remaining features, ALS patients were reclustered into two subgroups (Fig. 1d).

**Figure 1.**
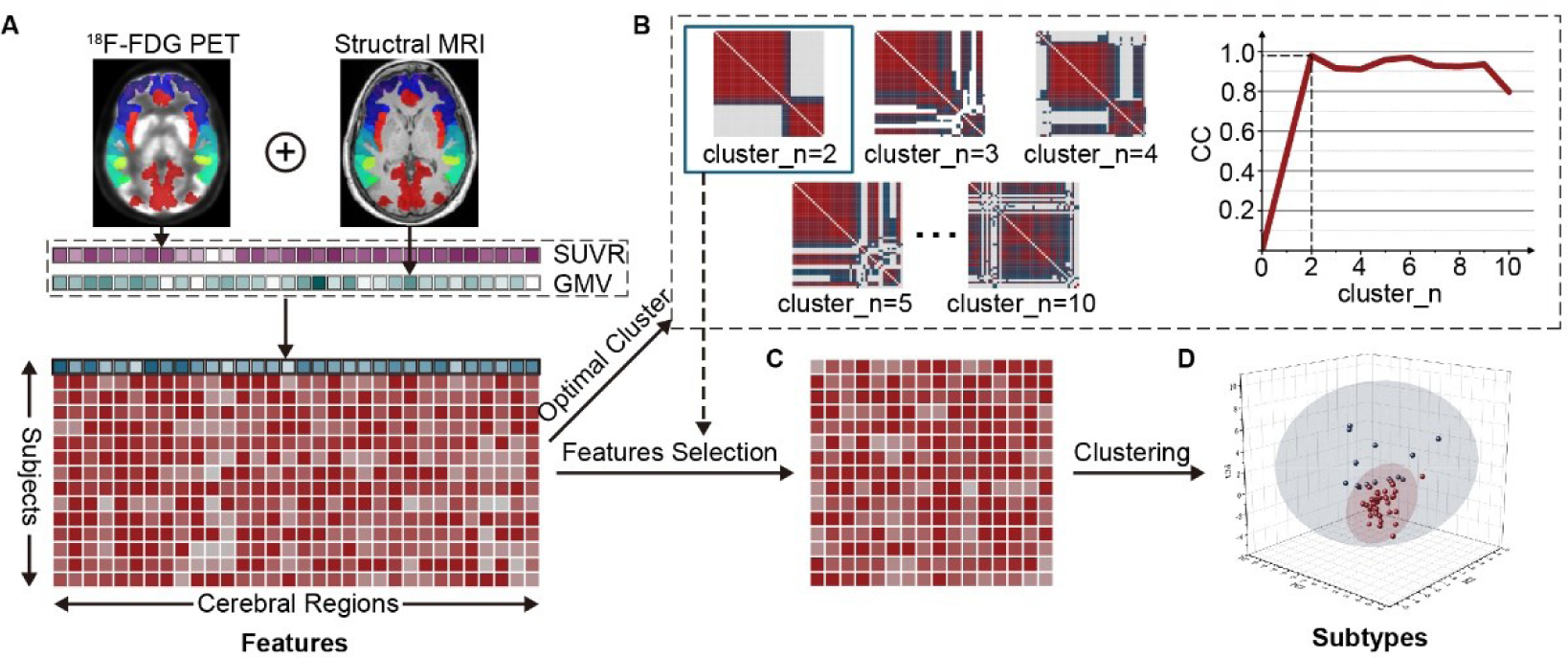
Cluster analysis procedure using ^18^F-FDG PET/MR imaging. A, Based on ^18^F-FDG PET and sMRI data, standardized uptake value ratio (SUVR) and gray matter volume (GMV) values in cerebral regions were extracted for every ALS patient and summed for each cerebral region to compose the features of the cluster analysis. B, The optimal number of clusters (cluster_n = 2) was obtained on account of the highest cophenetic correlation coefficient. C, With the optimal cluster, nonsignificant features by nonnegative matrix factorization (NMF) were deleted from the feature selection for the cluster analysis. D, Using the remaining features, ALS patients were reclustered into 2 subtypes, each displaying a spatially scattered distribution based on three principal components

### 2.6 Calculation of structural network measures

For brain WM network construction, the T1WI of each subject was aligned to the b0 image in native DTI space. Then, the aligned T1WI was transformed into the ICBM152 template in MNI space using FLRIT and FNIRT. The inverse transformation matrix was applied to warp the BNA246 and AAL90 from MNI space into native space. After the above procedures, we obtained two parcellations of each subject to separately define network nodes in native space. DTI tractography was performed through a deterministic tractography method with fractional anisotropy (FA) < 0.2 and angle > 45° as terminate parameters using the Diffusion Toolkit (https://www.trackvis.org/dtk/). An edge was defined if there was at least one streamline between two regions. The corresponding fiber number represents the weight of the edge. As a result, we constructed two fiber-number-weighted WM networks, which were two symmetric matrices of 246 × 246 and 90 × 90 for each subject. Measures of structural network, including the network global efficiency, network local efficiency and small-worldness (Lp, Cp, γ, λ and σ) at the global level as well as the nodal efficiency at the local level, were all derived by GRETNA software (http://www.nitrc.org/projects/gretna/) (Wang et al., 2015).

The NBS (Zalesky et al., 2010) approach was used to detect structural connection differences between groups from a subnetwork perspective. In particular, we first detected the significant nonzero connections within each group by statistical methods. A nonzero connection was defined as a connection present in more than half of the subjects in the group. Next, nonzero connections within the two compared groups were combined into a connection binary mask. The network of each participant was clipped by the Hadamard product with the binary mask. Finally, a toolbox (https://www.nitrc.org/projects/nbs) was used to identify the changed subnetworks in the context of pairwise comparisons. A primary threshold (*P* = 0.01 for ALS vs. HC; *P* = 0.05 for other group comparisons) was first applied to a two-sample one-tailed t test to compute a set of suprathreshold links. The components and number of these links were estimated for significance using a nonparametric permutation approach with 5,000 permutations. A value of *P* < 0.05 was considered significant, and Bonferroni corrections were used for group comparisons.

### 2.7 Support vector machine

Due to the regional characteristics of the structural network being sensitively represented by the nodal efficiency of the DTI-based brain network (dNE), we further aimed to create a new biomarker-based individualized network to recognize the clustered ALS subtypes. A support vector machine (SVM) classifier was trained to predict two clustered subtypes using the leave-one-out cross validation framework by a toolbox (https://www.csie.ntu.edu.tw/~cjlin/libsvm/), and an individualized network score (INS) was defined as the distance to the decision hyperplane in the feature space (Li et al., 2020). To this end, ALSFRS-R scores and DeltaFS were used to test whether INS was associated with clinical significance.

### 2.8 Statistical analyses

Differences in clinical features were compared by the Kruskal–Wallis test among the three groups and by the Wilcoxon rank-sum test between the two groups. Furthermore, data shown as percentages were compared in groups by chi-square tests.

For group comparisons of the global metrics of structural connectivity, dNE, GMV and SUVR, factors of age at PET/MR scan, sex ratio, and education years were previously removed by regression analysis. Next, the Kruskal–Wallis ANOVA test and subsequent post hoc pairwise comparisons were performed following pairwise comparisons. A value of *P* < 0.05 was considered significant. The false discovery rate (FDR) correction was used in the comparisons of dNE, GMV and SUVR. The Spearman correlation with FDR correction was applied between the significantly changed dNE and GMV or SUVR, while the Spearman correlation with Bonferroni correction was used between the INS and clinical characteristics. The Dice coefficient was used to analyze the percentage of labels with significantly different metrics in the cognition-related networks (Yeo et al., 2011).

## 3. Results

### 3.1 Clinical profiles of identified ALS subtypes

ALS patients and HCs were matched for age (Wilcoxon test, p = 0.73) and sex (chi-square test, p = 0.32). The identified two ALS subtypes were named locally impaired structural network (LISN) subtype and extensively impaired structural network (EISN) subtype. The demographic and clinical characteristics of the two ALS subgroups and HCs are summarized in Table 1.

**Table 1.**
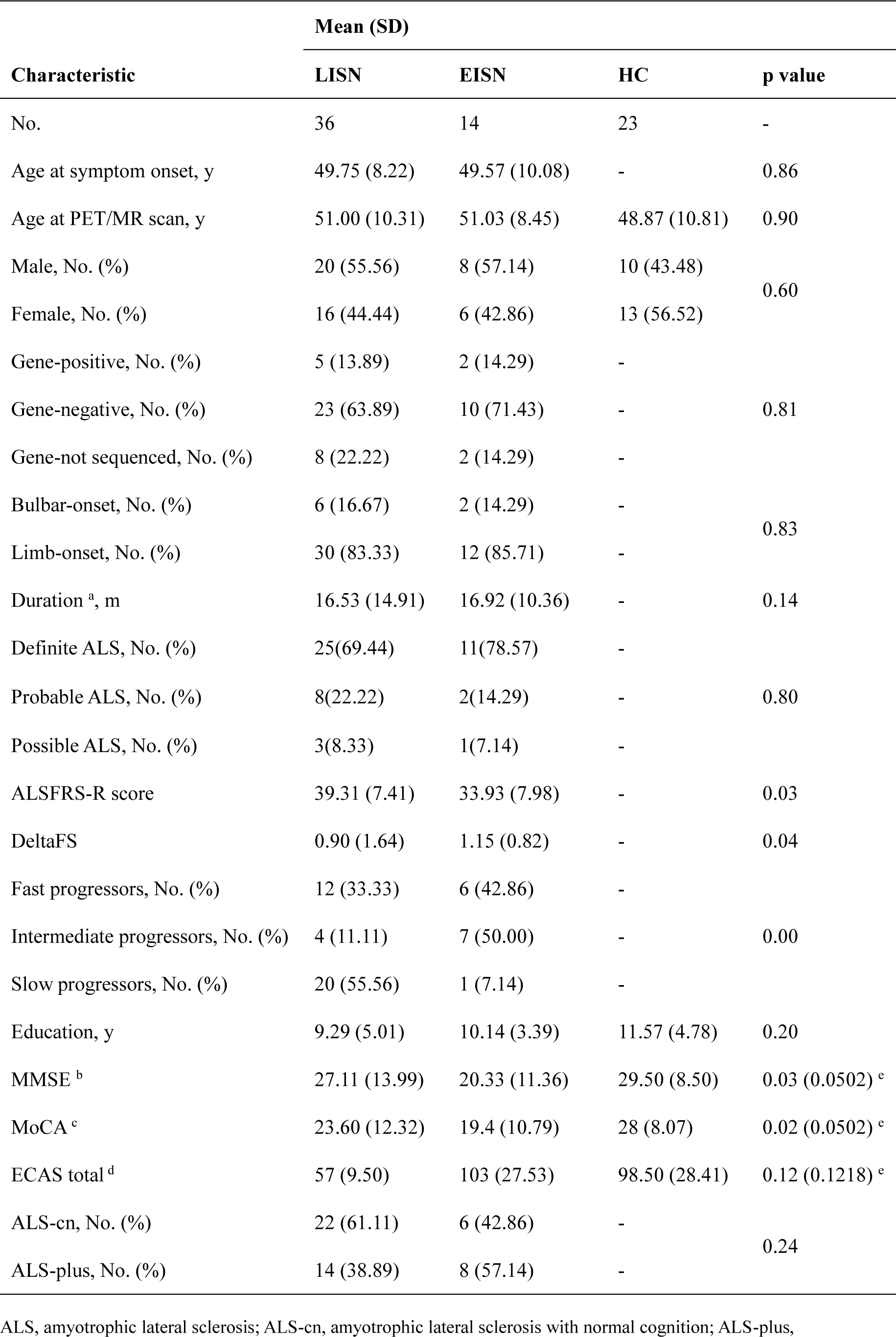
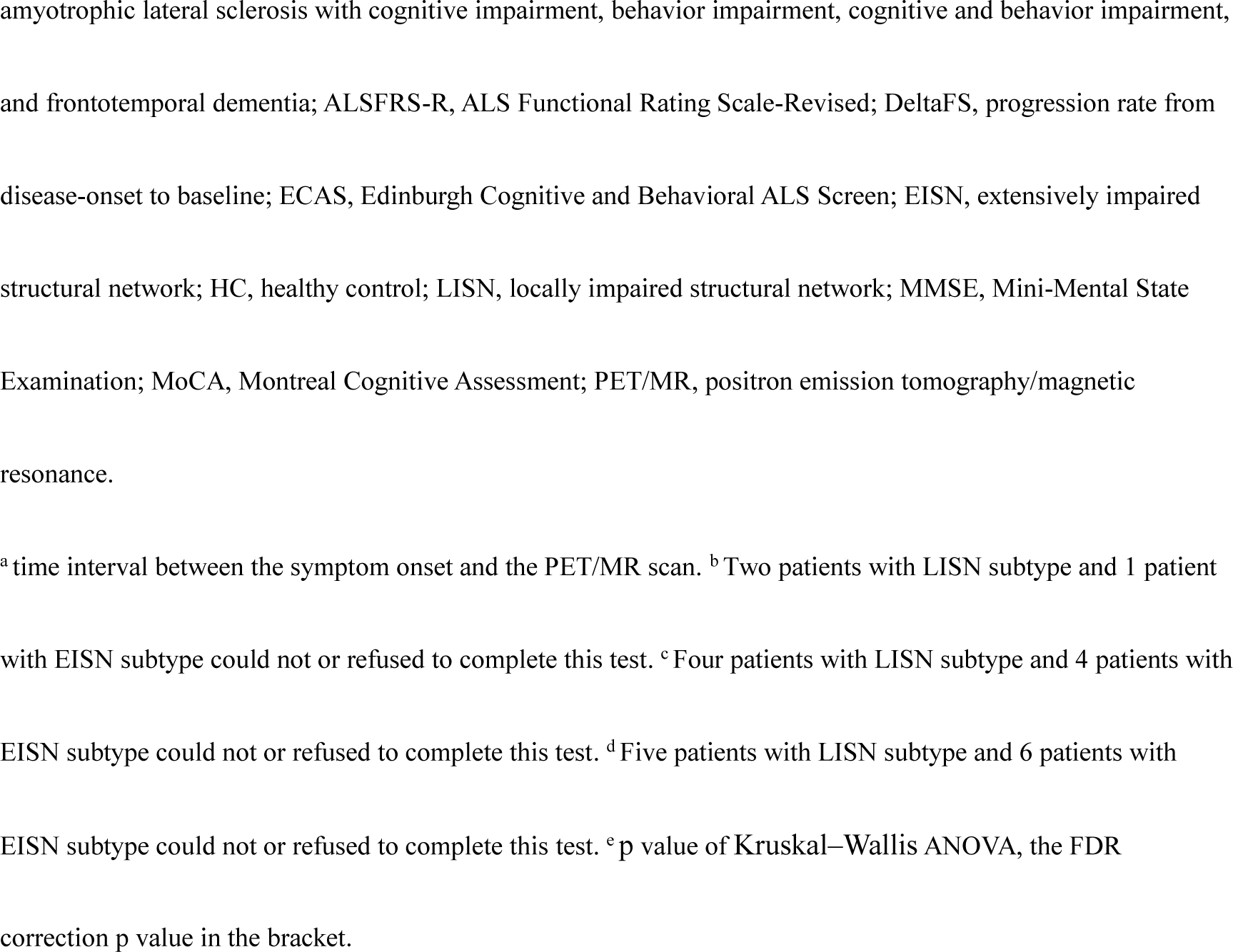
Demographic and clinical characteristics of participants.

| Characteristic | Mean (SD) |  |  | p value |
| --- | --- | --- | --- | --- |
|  | LISN | EISN | HC |  |
| No. | 36 | 14 | 23 | - |
| Age at symptom onset, y | 49.75 (8.22) | 49.57 (10.08) | - | 0.86 |
| Age at PET/MR scan, y | 51.00 (10.31) | 51.03 (8.45) | 48.87 (10.81) | 0.90 |
| Male, No. (%) | 20 (55.56) | 8 (57.14) | 10 (43.48) | 0.60 |
| Female, No. (%) | 16 (44.44) | 6 (42.86) | 13 (56.52) |  |
| Gene-positive, No. (%) | 5 (13.89) | 2 (14.29) | - | 0.81 |
| Gene-negative, No. (%) | 23 (63.89) | 10 (71.43) | - |  |
| Gene-not sequenced, No. (%) | 8 (22.22) | 2 (14.29) | - |  |
| Bulbar-onset, No. (%) | 6 (16.67) | 2 (14.29) | - | 0.83 |
| Limb-onset, No. (%) | 30 (83.33) | 12 (85.71) | - |  |
| Duration <sup>a</sup>, m | 16.53 (14.91) | 16.92 (10.36) | - | 0.14 |
| Definite ALS, No. (%) | 25(69.44) | 11(78.57) | - | 0.80 |
| Probable ALS, No. (%) | 8(22.22) | 2(14.29) | - |  |
| Possible ALS, No. (%) | 3(8.33) | 1(7.14) | - |  |
| ALSFRS-R score | 39.31 (7.41) | 33.93 (7.98) | - | 0.03 |
| DeltaFS | 0.90 (1.64) | 1.15 (0.82) | - | 0.04 |
| Fast progressors, No. (%) | 12 (33.33) | 6 (42.86) | - | 0.00 |
| Intermediate progressors, No. (%) | 4 (11.11) | 7 (50.00) | - |  |
| Slow progressors, No. (%) | 20 (55.56) | 1 (7.14) | - |  |
| Education, y | 9.29 (5.01) | 10.14 (3.39) | 11.57 (4.78) | 0.20 |
| MMSE <sup>b</sup> | 27.11 (13.99) | 20.33 (11.36) | 29.50 (8.50) | 0.03 (0.0502) <sup>c</sup> |
| MoCA <sup>c</sup> | 23.60 (12.32) | 19.4 (10.79) | 28 (8.07) | 0.02 (0.0502) <sup>c</sup> |
| ECAS total <sup>d</sup> | 57 (9.50) | 103 (27.53) | 98.50 (28.41) | 0.12 (0.1218) <sup>c</sup> |
| ALS-cn, No. (%) | 22 (61.11) | 6 (42.86) | - | 0.24 |
| ALS-plus, No. (%) | 14 (38.89) | 8 (57.14) | - |  |

ALS, amyotrophic lateral sclerosis; ALS-cn, amyotrophic lateral sclerosis with normal cognition; ALS-plus,
amyotrophic lateral sclerosis with cognitive impairment, behavior impairment, cognitive and behavior impairment, and frontotemporal dementia; ALSFRS-R, ALS Functional Rating Scale-Revised; DeltaFS, progression rate from disease-onset to baseline; ECAS, Edinburgh Cognitive and Behavioral ALS Screen; EISN, extensively impaired structural network; HC, healthy control; LISN, locally impaired structural network; MMSE, Mini-Mental State Examination; MoCA, Montreal Cognitive Assessment; PET/MR, positron emission tomography/magnetic resonance.
<sup>a</sup>time interval between the symptom onset and the PET/MR scan. <sup>b</sup>Two patients with LISN subtype and 1 patient with EISN subtype could not or refused to complete this test. <sup>c</sup>Four patients with LISN subtype and 4 patients with EISN subtype could not or refused to complete this test. <sup>d</sup>Five patients with LISN subtype and 6 patients with EISN subtype could not or refused to complete this test. <sup>e</sup>p value of Kruskal–Wallis ANOVA, the FDR correction p value in the bracket.

No difference was found between patients with LISN and EISN subtype and HCs either in age at symptom onset (49.75 ± 8.22 vs. 49.57 ±10.08, p = 0.86), age at PET/MR scan (51.00 ± 10.31 vs. 51.03 ± 8.45 vs. 48.87 ± 10.81, p = 0.90), education (9.29 ± 5.01 vs. 10.14 ± 3.39 vs. 11.57 ± 4.78, p = 0.20) and duration (16.53 ± 14.91 vs. 16.92 ± 10.36, p = 0.14), or in the distribution of gender (16 F (44.44 %) vs. 6 F (42.86 %) vs. 13 F (56.52 %), p = 0.60), gene (p = 0.81), site of onset (30 Limb-onset (83.33 %) vs. 12 Limb-onset (85.71 %), p = 0.83) and the percent of patients with definite ALS, probable ALS and possible ALS (p = 0.80). Notably, the LISN subgroup showed significantly higher ALSFRS-R scores and slower DeltaFS than the EISN subgroup (p < 0.05).

The scores of MMSE, MoCA and ECAS total were found no significant differences in subgroups comparisons (p > 0.05, FDR correction). The ECAS subscores of three subgroups were displayed in Table 2. None of them showed significant difference in subgroups comparisons (p > 0.05, FDR correction). Furthermore, percent of patients with ALS-cn and with ALS-plus showed no significant difference in the comparison of LISN and EISN subgroups (p > 0.05).

**Table 2.**
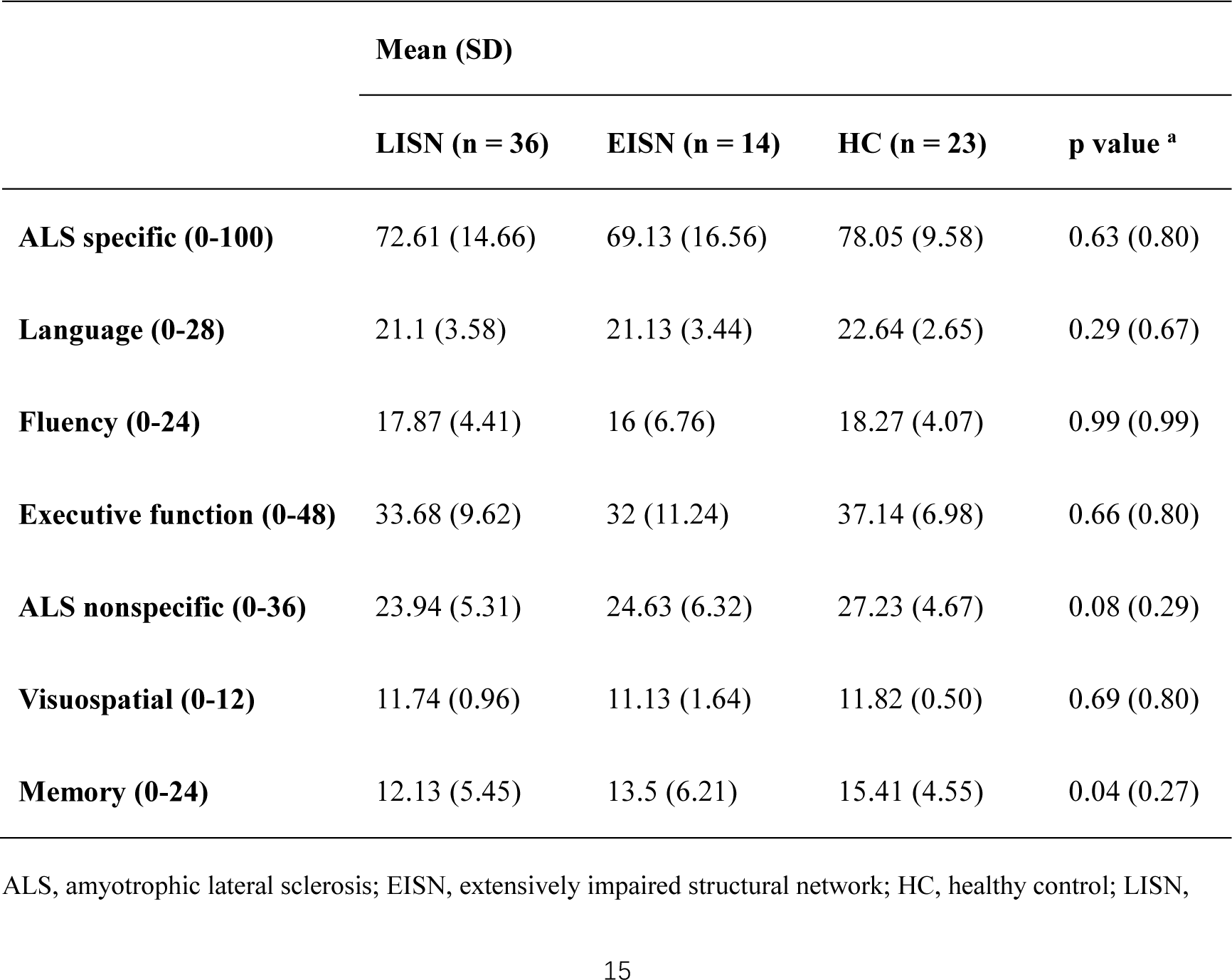
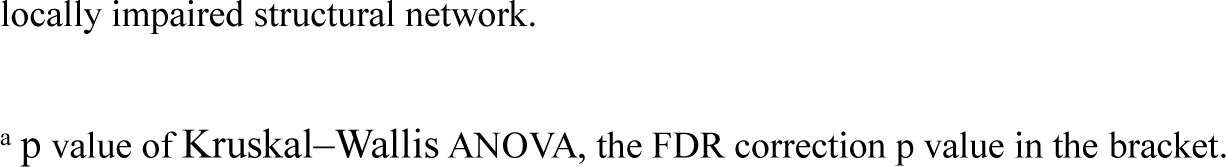
Group comparisons of ECAS subscores of participants.

|  | Mean (SD) |  |  |  |
| --- | --- | --- | --- | --- |
|  | LISN (n = 36) | EISN (n = 14) | HC (n = 23) | p value <sup>a</sup> |
| **ALS specific (0-100)** | 72.61 (14.66) | 69.13 (16.56) | 78.05 (9.58) | 0.63 (0.80) |
| **Language (0-28)** | 21.1 (3.58) | 21.13 (3.44) | 22.64 (2.65) | 0.29 (0.67) |
| **Fluency (0-24)** | 17.87 (4.41) | 16 (6.76) | 18.27 (4.07) | 0.99 (0.99) |
| **Executive function (0-48)** | 33.68 (9.62) | 32 (11.24) | 37.14 (6.98) | 0.66 (0.80) |
| **ALS nonspecific (0-36)** | 23.94 (5.31) | 24.63 (6.32) | 27.23 (4.67) | 0.08 (0.29) |
| **Visuospatial (0-12)** | 11.74 (0.96) | 11.13 (1.64) | 11.82 (0.50) | 0.69 (0.80) |
| **Memory (0-24)** | 12.13 (5.45) | 13.5 (6.21) | 15.41 (4.55) | 0.04 (0.27) |

ALS, amyotrophic lateral sclerosis; EISN, extensively impaired structural network; HC, healthy control; LISN,
locally impaired structural network.
<sup>a</sup> p value of Kruskal–Wallis ANOVA, the FDR correction p value in the bracket.

### 3.2 Alterations of structural network

Globally, the small-worldness (Lp, Cp, γ, λ and σ) showed no significant difference in all group comparisons (p > 0.05). For the network global and local efficiency, there were significant differences (p < 0.05, FDR correction) (Table 3) in the comparisons of LISN, EISN and HC groups. Compared with HC, the network global and local efficiency in the LISN subgroup showed no significant changes (p > 0.05), and that in the EISN subgroup were significantly lower (p < 0.05) (Table 3) but presented no correlations with ALSFRS-R scores and DeltaFS (p > 0.05). In the comparisons of ALS and HC groups, the network global and local efficiency were both not significantly different (p > 0.05, FDR correction) (Table 3).

**Table 3.** Group comparisons of global measures of structural network with BNA.

|  | LISN, EISN,<br>vs. HC | LISN vs. HC |  | EISN vs. HC |  | EISN vs. LISN |  | ALS vs. HC |  |
| --- | --- | --- | --- | --- | --- | --- | --- | --- | --- |
|  |  | p | t | p | t | p | t | p | t |
| NGE | 0.00 (0.02)<sup>a</sup> | 0.08 | 2.14 | 0.00<sup>b</sup> | 3.34 | 0.78 | 1.78 | 0.02 (0.15)<sup>c</sup> | -2.34 |
| NLE | 0.01 (0.047)<sup>a</sup> | 0.18 | 1.76 | 0.01<sup>b</sup> | 2.91 | 0.23 | 1.64 | 0.046 (0.16)<sup>c</sup> | -2.03 |

ALS, amyotrophic lateral sclerosis; BNA, Brainnetome Atlas; EISN, extensively impaired structural network; HC, healthy control; LISN, locally impaired structural network; NGE, network global efficiency; NLE, network local efficiency.
<sup>a</sup> p value of Kruskal–Wallis ANOVA, the FDR correction p value in the bracket. <sup>b</sup> Values of measures in the latter group are higher than that in the former group. <sup>c</sup> p value of Wilcoxon rank-sum test, the FDR correction p value in the bracket.

Locally, compared with HC, labels with decreases in dNE exhibited a distribution pattern centralized in the sensorimotor network in the LISN subgroup but a widespread involvement of the frontal, parietal and temporal lobes as well as subcortical regions in the EISN subgroup (p < 0.05, FDR correction) (Fig. 2a; Table S2). Labels with increases in dNE were absent in all group comparisons.

**Figure 2.**
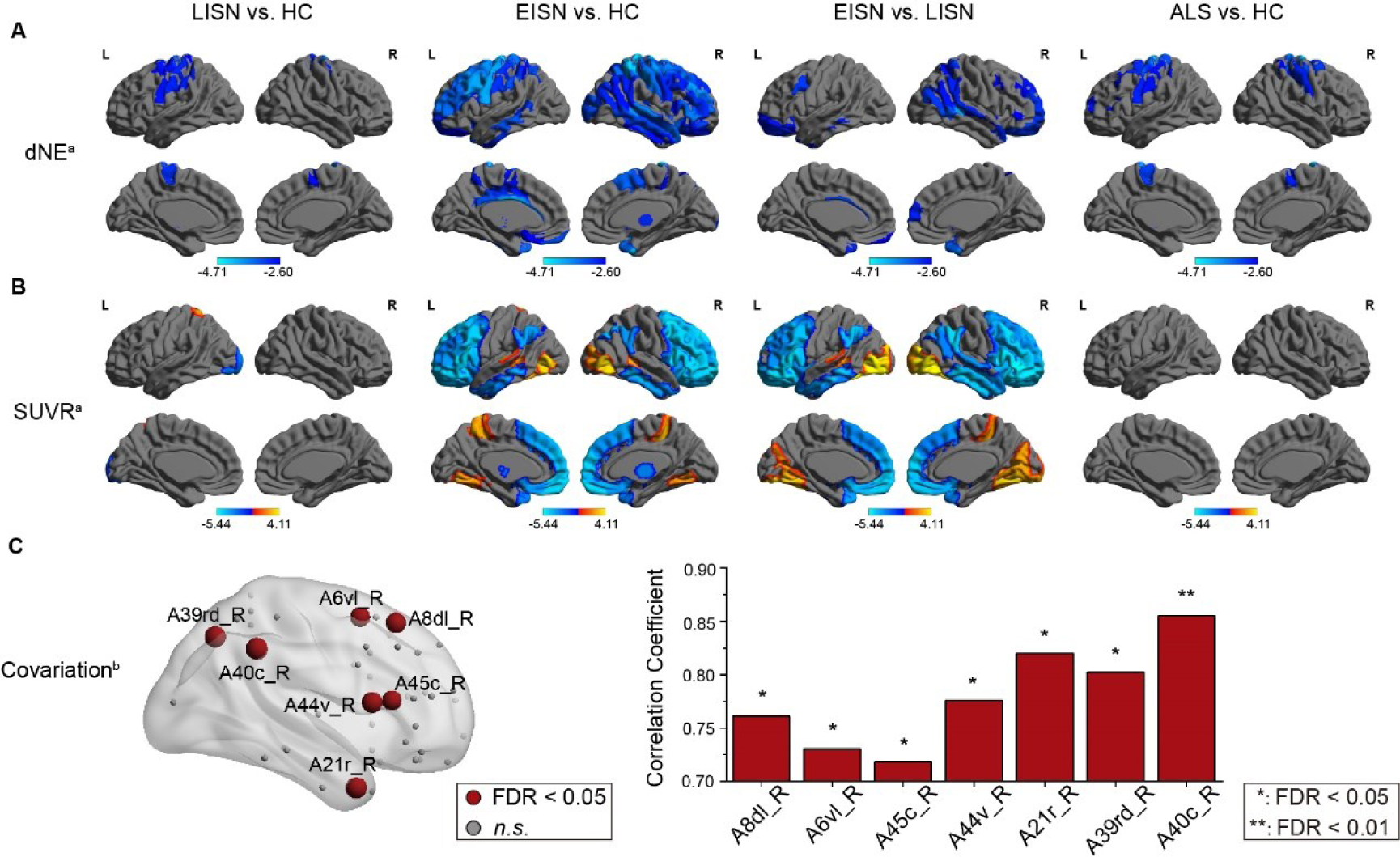
Distribution patterns of labels with decreases in dNE and changes in SUVR with the BNA. A, Distribution pattern of labels with decreases in dNE. Color represent *P* values of the labels. B, Distribution pattern of labels with decreases and increases in SUVR. Color represent *P* values of the labels. C, Labels with related decreases in dNE and SUVR in the EISN subgroup. The volumes of the red balls represent the Spearman r values of the labels. ALS indicates amyotrophic lateral sclerosis; dNE, nodal efficiency of the DTI-based brain network; EISN, extensively impaired structural network; HC, healthy control; LISN, locally impaired structural network; SUVR, standardized uptake value ratio. ^a^ p < 0.05, FDR correction. ^b^ Labels with related decreases in dNE and SUVR in the EISN subgroup

NBS identified different structural subnetworks with significant decreases in connections when comparing the LISN subgroup and EISN subgroup with HC (p < 0.05, Bonferroni correction) (Fig. 3). The impaired structural subnetwork in the LISN vs. HC comparison mainly contained links interconnecting the bilateral precentral gyri and postcentral gyri with the subcortical regions (Fig. 3a), while that in the EISN vs. HC comparison displayed a complicated composition with links interconnecting the widespread cerebral regions (Fig. 3b). Furthermore, in the EISN vs. LISN comparison, 3 impaired structural subnetworks were observed (Fig. 3c), which included connections within the bilateral frontal regions in component 1, connections in the right hemisphere in component 2, and connections in the left hemisphere in component 3.

**Figure 3.**
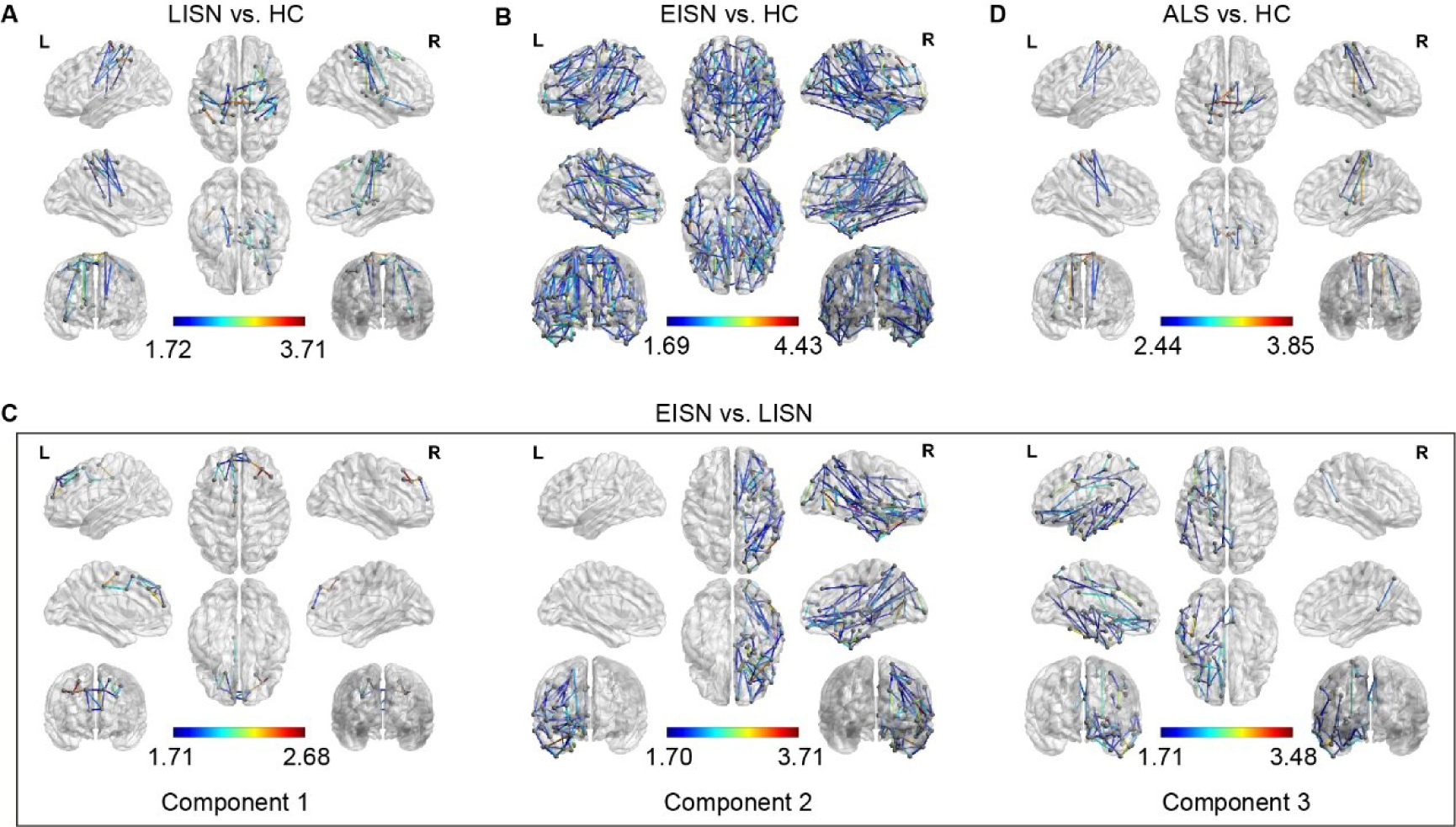
The impaired structural subnetworks in group comparisons based on the BNA. The impaired structural subnetworks in LISN vs. HC (A), EISN vs. HC (B), EISN vs. LISN (C), and ALS vs. HC (D) comparisons (p < 0.05, Bonferroni correction) by network-based statistics (NBS). Color represent p values of the edges constituting the impaired subnetworks. ALS indicates amyotrophic lateral sclerosis; EISN, extensively impaired structural network; HC, healthy control; LISN, locally impaired structural network

### 3.3 GMV and SUVR changes

GMV of cerebral regions all showed no significant changes in the LISN vs. HC, EISN vs. HC, EISN vs. LISN and ALS vs. HC comparisons (p > 0.05, FDR correction). Compared with HC, ^18^F-FDG hypometabolism was only distributed in the left lateral occipital cortex in the LISN subgroup but was widely distributed in the bilateral orbital gyri and superior frontal gyri, left inferior frontal gyrus, and right middle frontal gyrus in the EISN subgroup. In addition, patients with the LISN subtype showed ^18^F-FDG hypermetabolism only in the left superior parietal lobule, while patients with the EISN subtype displayed ^18^F-FDG hypermetabolism in the bilateral lateral occipital cortices and precuneus and left fusiform gyrus, paracentral lobule, superior parietal lobule and thalamus when compared with HCs (p < 0.05, FDR correction) (Fig. 2b; Table S3). Notably, although 18 patients with ALS-cn showed normal ^18^F-FDG metabolism in the general observation, 5 of them were clustered into the EISN subgroup. Among the 10 ALS-cn patients with ^18^F-FDG hypometabolism in the general observation, 9 patients were clustered into the LISN subgroup (Table S4).

### 3.4 Decreases in dNE and SUVR

In the EISN subgroup, labels with positively related decreases in dNE and SUVR (p < 0.05, FDR correction) were demonstrated in the right superior frontal gyrus, middle frontal gyrus, inferior frontal gyrus, middle temporal gyrus, and inferior parietal lobule (Fig. 2c; Table S5).

As Table S6 shows, the percentage of labels with decreases in dNE and SUVR in the cognition-related networks (Yeo et al., 2011) was zero or low in the LISN vs. HC comparison and was high in many of the cognition-related networks in the EISN vs. HC comparison. In particular, the percentage of labels with decreases in dNE was over half in the somatomotor network, dorsal attention network, ventral attention network, frontoparietal network and default network, while the percentage of labels with decreases in SUVR was more than half in the limbic network, frontoparietal network and default network when comparing the EISN subgroup to HC.

### 3.5 INS in patients with ALS

As Fig. 4 shows, the INS in every ALS patient was obtained by an SVM classifier based on the decreases in dNE in the EISN vs. LISN comparison. In all ALS patients, a positive correlation was found between the INS and ALSFRS-R scores (r = 0.37, p < 0.05, Bonferroni correction), and a negative correlation was observed between the INS and DeltaFS (r = −0.44, p < 0.05, Bonferroni correction).

**Figure 4.**
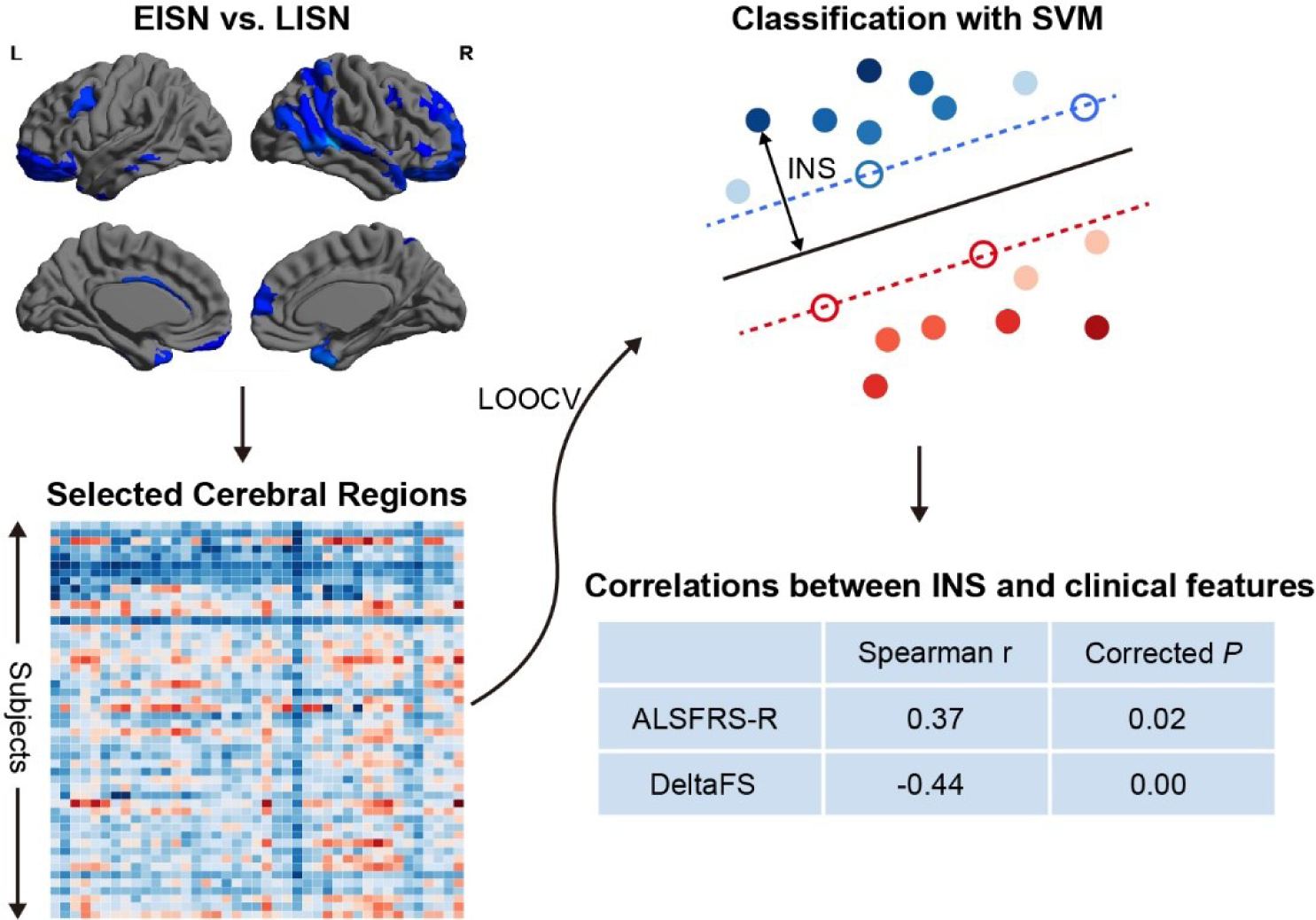
INS for ALS patients using SVM. One support vector machine (SVM) classifier by dNE was trained to identify EISN and LISN using the leave-one-out cross validation framework. Individualized network scores (INS) were defined as the distance to the decision hyperplane in the feature space. A positive correlation between INS and ALSFRS-R scores and a negative correlation between INS and disease progression rate were revealed. ALS indicates amyotrophic lateral sclerosis; ALSFRS-R, ALS Functional Rating Scale-Revised; DeltaFS, progression rate from disease-onset to baseline; EISN, extensively impaired structural network; LISN, locally impaired structural network; LOOCV, leave-one-out cross validation

### 3.6 Validation with the AAL

The distinct patterns of structural network impairments in clustered subtypes based on the BNA were similarly validated with the AAL. With the AAL, the small-worldness (Lp, Cp, γ, λ and σ) showed no significant difference in all group comparisons (p > 0.05). Compared with HC, the network global and local efficiency in the LISN subgroup showed no significant changes (p > 0.05), and that in the EISN subgroup was significantly lower (p < 0.05) (Table S7) but presented no correlations with ALSFRS-R scores and DeltaFS (p > 0.05). Compared with HC, labels with decreases in dNE exhibited the similar distribution patterns in the LISN and EISN subgroups (p < 0.05, FDR correction) (Fig. S1a). Labels with increases in dNE were absent in all group comparisons. NBS displayed an impaired structural subnetwork involving connections in the sensorimotor network in the LISN vs. HC comparison (Fig. S2a), and an impaired structural subnetwork with links interconnecting the widespread cerebral regions (Fig. S2b).

Similarly, GMV of cerebral regions showed no significant reductions in all group comparisons (p > 0.05, FDR correction). Compared with HC, ^18^F-FDG hypometabolism and hypermetabolism were all absent in the LISN subgroup but exhibited a similar distribution pattern in the EISN subgroup (p < 0.05, FDR correction) (Fig. S1b). Labels with positively related decreases in dNE and SUVR (p < 0.05, FDR correction) were mainly observed in the bilateral superior frontal gyrus, right middle frontal gyrus and inferior frontal gyrus in the EISN subgroup (Fig. S1c; Table S8). The percentages of labels with decreases in dNE and SUVR in the cognition-related networks are shown in Table S6 and are similar to those based on the BNA.

## 4. Discussion

By cluster analysis based on GMV and SUVR, we identified two ALS subtypes as the optimal cluster, the LISN subtype and EISN subtype. Because the two ALS subtypes are identified by a data-driven method, we speculate that ALS possesses an intrinsic pattern of WM damages.

Compared to HC, the network global and local efficiency were found to be lower in the EISN subgroup but not in the LISN subgroup. The EISN subgroup exhibited global network alterations, which is in line with the reports of decreases in global efficiency (Fortanier et al., 2019; Zhang et al., 2019) and local efficiency (Basaia et al., 2020) in ALS patients compared to controls. However, previous studies also found no significant differences in network efficiency between the ALS and HC groups (Buchanan et al., 2015; Dimond et al., 2017). The discrepancy may be due to the different patient inclusion criteria and in support of the divergence of structural network impairments found in our study. In addition, the network global and local efficiency represent the integration and segregation ability of information transfer. Thus, the clustered EISN subtype is supposed to encompass global dysconnectivity in WM networks.

At the regional scale, decreases in dNE were centralized in the sensorimotor network in the ALS vs. HC and LISN vs. HC comparisons, while exhibited a widespread distribution in the EISN vs. HC comparison. The network efficiency of one node quantifies the efficiency of parallel information transfer by that node in the network. Thus, our findings showed that the declined ability of information transfer in the LISN subtype was limited within the sensorimotor network but was widespread in almost the whole brain in the EISN subtype. Previously, degeneration of the sensorimotor network has been reported in ALS patients by widespread precentral and postcentral FA reductions (Rose et al., 2012). Thus, the decreased nodal efficiency in the sensorimotor network may result from the disruption of WM integrity. In addition, decreases in dNE in the bilateral frontal and temporal cortexes, right gyrus rectus, paracentral lobule and caudate were also reported in ALS patients compared to HCs (Li et al., 2021). Our results also demonstrated decreases in dNE in regions beyond the sensorimotor network in ALS patients. Furthermore, in the EISN vs. HC comparison, a few subregions of the right frontal, parietal and temporal cortices displayed positively correlated decreases in dNE and ^18^F-FDG hypometabolism. The decline in dNE represents the reduced efficiency of information transfer and is supposed to produce the correlated ^18^F-FDG hypometabolism in the impaired brain regions.

Our findings by NBS support the view that WM changes make up subnetwork of impaired connectivity and further uncover the diversely impaired structural subnetwork in the clustered ALS subtypes. The composition of the impaired structural subnetwork in the LISN subgroup was highly consistent with the previously reported impaired motor subnetwork centered on the precentral and paracentral nodes (Basaia et al., 2020; Fortanier et al., 2019) when compared with HC. However, alterations involving the connections within and among the sensorimotor network, basal ganglia, frontal, temporal, and parietal areas were found in ALS-cn, ALS-ci/bi and ALS-FTD patients but with a more widespread disruption in ALS-FTD patients when compared to controls (Cividini et al., 2021). The largest connected component in ALS-cn patients was centralized around the motor system, while that in ALS-ci patients included frontal and temporal connections and that in ALS-bi patients included motor, temporal, frontal, and parietal connections (van der Burgh et al., 2020). Thus, the impaired structural subnetwork with extensive connections in the whole brain in the EISN subgroup implied the damages of cognition. For ALS patients, neuropsychological tests are the preliminary selection for the evaluation of cognitive and behavioral levels by clinicians. Those patients with abnormal neuropsychological assessments should accept a further ^18^F-FDG PET scan, which provides more clues for the evaluation of cognitive and behavioral impairment. Normal neuropsychological evaluation and ^18^F-FDG metabolism generally imply no cognitive and behavioral impairment in ALS patients. However, in our cohort, 5 ALS-cn patients with normal ^18^F-FDG metabolism in the general observation were clustered into the EISN subgroup, while 9 ALS-cn patients with ^18^F-FDG hypometabolism in the general observation were clustered into the LISN subgroup. As we know, the neuropsychological evaluation inevitably contains subjectivity from ALS patients and their relatives or caregivers and could be affected or limited by the dysarthria and dysfunction of upper limbs in ALS patients. The results of ^18^F-FDG metabolism in the general observation may encompass some errors. Thus, compared with the categorization based on the clinical features, our cluster analysis reflects clues for cognitive assessment in ALS patients from a data-driven perspective. Due to the remarkable percentage of labels with decreases in dNE and ^18^F-FDG hypometabolism in cognition-related networks (Yeo et al., 2011) in the EISN subgroup, the clustered EISN patients with ALS-cn and normal ^18^F-FDG metabolism are expected to have risks of developing cognitive and behavioral impairment. Correspondingly, the clustered LISN patients with ALS-cn and ^18^F-FDG hypometabolism in the general observation are supposed to have unimpaired cognition, according to the low percentage of labels with ^18^F-FDG hypometabolism and the decreases in dNE in the cognition-related networks in the LISN subgroup.

Based on the decreases in dNE in the EISN vs. LISN comparison, we constructed a classifier to obtain an INS for every ALS patient. The positive relation between INS and ALSFRS-R scores and the negative relation between INS and disease progression rate in all ALS patients, indicated that more decreases in dNE indicated more severe disease. Thus, the decreases in dNE may be a potential biomarker for the phenotypes of ALS. Furthermore, ALS has no effective treatment or cure thus far, which may result from the high heterogeneity of clinical features. Based on the clustered subtypes by a data-driven method and the decreases in dNE related to disease severity, our findings could contribute to a latent direction for stratified research about medicine or remedy in the future.

This study has limitations. First, the sample size was relatively small, but the results are encouraging and deserve further investigation in a larger cohort as well as validation in another independent cohort. Second, the structural networks were constructed by an atlas-based pipeline, not by a high-resolution vertex-level pipeline that may encompass the potential advantages. However, we have made a validation with the AAL. Finally, as a cross-sectional observation, the follow-up of clinical features and PET/MR examination were expected to uncover the progression of clustered subtypes.

## 5. Conclusions

We demonstrate for the first time that the subtypes of ALS patients can be clustered by a data-driven analysis using PET/MR hybrid data. The two subtypes identified as the optimal cluster encompass different patterns of structural network impairments. The demonstration that decreases in dNE are correlated with disease severity implies a new possibility in the selection of biomarkers for the phenotypes of ALS. Our findings can provide objective information for ALS, thus facilitating clinical evaluation and providing the latent direction for stratified therapies.

### Ethics approval and consent to participate

The study was approved by the Medical Ethics Committee of the Chinese PLA General Hospital, Beijing, China (S2020-027-01). All participates provided a written informed consent.

### Data Availability

The datasets used during the current study are available from the corresponding authors on reasonable request.

### Funding

This work was supported by the National Natural Science Foundation of China (No. 81671761 and 81871425); Fundamental Research Funds for the Central Universities (No. 2017XTCX04); Open Research Fund of the State Key Laboratory of Cognitive Neuroscience and Learning (No. CNLYB2001).

### Author contributions

**Feng Feng:** Investigation, Resources, Formal analysis, Writing - Original Draft. **Guozheng Feng:** Resources, Formal analysis, Software, Writing - Original Draft. **Jiajin Liu:** Investigation, Resources, Writing - Original Draft. **Weijun Hao:** Investigation. **Weijie Huang:** Investigation. **Xiao Bi:** Investigation. **Mao Li:** Investigation. **Hongfen Wang:** Investigation. **Fei Yang:** Investigation. **Zhengqing He:** Investigation. **Jiongming Bai:** Investigation. **Haoran Wang:** Investigation. **Guolin Ma:** Investigation. **Baixuan Xu:** Conceptualization, Methodology, Writing - Review & Editing, Supervision. **Ni Shu:** Conceptualization, Methodology, Writing - Review & Editing, Funding acquisition, Supervision. **Xusheng Huang:** Conceptualization, Methodology, Writing - Review & Editing, Supervision.

## Abbreviations

AAL: Automated anatomical labeling
ALS: Amyotrophic lateral sclerosis
ALS-bi: ALS with behavioral impairment
ALS-cbi: ALS with cognitive and behavioral impairment
ALS-ci: ALS with cognitive impairment
ALSFRS-R: ALS Functional Rating Scale-Revised
BNA: Brainnetome Atlas
DeltaFS: Progression rate from disease-onset to baseline
dNE: Nodal efficiency of DTI-based brain network
EISN: Extensively impaired structural network
FTD: Frontotemporal dementia
GMV: Gray matter volume
HC: Healthy control
INS: Individualized network score
LISN: Locally impaired structural network
LOOCV: Leave-one-out cross validation
NBS: Network-based statistics
sMRI: Structural MRI
SUVR: Standardized uptake value ratio
SVM: Support vector machine
WM: White matter

## Acknowledgements

We thank all participants and their relatives for their contributions to this research.

